# The impact of COVID vaccination on incidence of long COVID and healthcare resource utilisation in a primary care cohort in England, 2021-2022

**DOI:** 10.1101/2024.04.24.24306308

**Authors:** Jingyan Yang, Kiran K. Rai, Tamuno Alfred, Lucy Massey, Olivia Massey, Leah McGrath, Kathleen M. Andersen, Theo Tritton, Carmen Tsang, Rebecca Butfield, Charlie Reynard, Diana Mendes, Jennifer L Nguyen

## Abstract

**Background:** Long COVID, a diverse set of symptoms that persist after a minimum of 4 weeks from the initial SARS-CoV-2 infection, has posed substantial burden to healthcare systems. There is some evidence that COVID-19 vaccination may be associated with lower risk of long COVID. However, little is known about the association between vaccination status and long COVID-associated healthcare resource utilisation (HCRU) and costs.

**Methods:** We conducted a cohort study using primary care electronic health record data in England from the Clinical Practice Research Datalink (CPRD) Aurum dataset linked to Hospital Episode Statistics where applicable. Adult (≥18 years) patients were indexed on a COVID-19 diagnosis between 1^st^ March 2021 and 1^st^ December 2021. Vaccination status was assessed at index: unvaccinated or completed primary series (two doses for immunocompetent and three doses for immunocompromised patients). Covariate balance was conducted using entropy balancing. Weighted multivariable Poisson regression was used to estimate the incidence rate ratio (IRR) for incident long COVID, and separately long COVID primary care resource use, by vaccination status. Patients were followed up to a maximum of 9-months post index.

**Results:** A total of 35,713 patients who had completed primary series vaccination, and 75,522 unvaccinated patients were included. The weighted and adjusted IRR for long COVID among patients vaccinated with the primary series compared to being unvaccinated was 0.81 (95% CI: 0.77-0.86) in the overall cohort, 0.83 (95% CI: 0.78-0.88) in the immunocompetent cohort and 0.28 (95% CI: 0.13-0.58) in the immunocompromised cohort. Among those with long COVID, there was no association between the rate of primary care consultations and vaccination status in the overall and immunocompetent cohorts. Cost of primary care consultations was greater in the unvaccinated group than for those who completed primary series.

**Conclusion:** Vaccination against COVID-19 may reduce the risk of long COVID in both immunocompetent and immunocompromised patients. However, no association was found between frequency of primary care visits and vaccination among patients diagnosed in 2021. Future studies with larger sample size, higher vaccine uptake, and longer study periods during the pandemic are needed to further quantify the impact of vaccination on long COVID.

## Background

As of December 2023, coronavirus disease 2019 (COVID-19) has led to more than 21 million cases, more than 1 million hospitalisations and approximately 197,000 deaths in the UK [1]. Symptoms of infection may persist for months, and in October 2021, the World Health Organization (WHO) released a clinical case definition for post-COVID-19 condition as symptoms that are present 3 months after SARS-CoV-2 infection with a minimum duration of 2 months which cannot be explained by alternative diagnosis [2]. These symptoms are collectively and commonly referred to as long COVID. The manifested symptoms are heterogeneous, including fatigue, fever/chills, brain fog, and shortness of breath, and may be experienced for varying time periods [3, 4]. Long COVID impacts on multiple organ systems, with potentially several hypotheses for its pathogenesis including immune dysregulation, microbiota disruption, blood clotting and endothelial abnormalities, autoimmunity and dysfunctional neurological signalling [5]. Data from the UK’s Office for National Statistics show that, as of 5^th^ March 2023, long COVID affected approximately 3.0% of the population [6], although a Scottish cohort study has reported higher adjusted estimates ranging from 6.6% to 10.4% for varying times since the initial infection [3]. The primary care costs (GP, nurse and physiotherapy visits) in non-hospitalised adults with long COVID in the UK are substantial and were estimated to cost the UK economy £23.4 million between May 2020 and April 2021 [7]. Further, costs may be much higher as medication costs and other community care support have not been factored into these estimates. There is also some evidence showing long COVID is associated with poorer work productivity and quality of life, even among those who experienced mild-to-moderate infection [8-11].

In the UK, more than 75% of the population had received at least two doses of COVID vaccine by September 2021 [12]. Emerging evidence has suggested protective effects of vaccination on long COVID. A systematic review of observational studies (case control and cohort studies) demonstrated that COVID-19 vaccination prior to infection was significantly associated with reduced risk of long COVID [13]. Further, data from a French prospective cohort study demonstrated that among those with long COVID, vaccination was associated with reduced symptom severity [14]. However, the following limitations have been noted: limited evidence arising from UK data; long COVID definitions based on self-report [15] or using the WHO definition for symptoms at the time of the study, which included fewer symptoms than more recent research [3, 16]. Separately, whilst there is evidence describing the economic burden of long COVID, little is known about the impact of vaccination on primary care resource use in long COVID patients. We therefore conducted a retrospective study using data from a large UK primary care database to assess among patients who had been infected with pre-Omicron SARS-CoV-2 strains 1) the incidence of long COVID and 2) health care resource utilisation (HCRU) and costs among patients with long COVID, compared between patients who had completed primary series vaccination and unvaccinated patients,

## METHODS

### Study design and setting

We conducted a population-based retrospective cohort study using primary care data from the Clinical Practice Research Datalink (CPRD-Aurum) and linked secondary care administrative data from Hospital Episode Statistics, Admitted Patient Care dataset (HES APC) where available. The May 2022 release of CPRD Aurum was used. The study design and methods have been described elsewhere [17]. See **Supplementary figure 1** for the study design schematic.

### Population

Adult patients (aged ≥18 years) diagnosed with COVID-19 between 1^st^ March 2021 and 31^st^ March 2021 that did not have a record for a COVID-19 related hospitalisation (i.e. those with a COVID-19 hospitalisation during March 2021 were not included) *as well as* all persons diagnosed on or after 1^st^ April 2021 to 1^st^ December 2021 (the period of time for which CPRD did not have hospitalisation data available) were included in this cohort. The index period start date was chosen to align with the date of availability of the complete COVID-19 vaccine primary series, whereas the end date was aligned to data availability and the period prior to the dominance of the Omicron variant in the UK.

### Exposure

Details on COVID-19 vaccination definitions have been previously reported [17]. In brief, product and medical codes, regardless of brand, were considered. An immunocompetent patient was considered vaccinated starting from 14 days after receipt of dose 2, and each dose was required to be separated by at least 21 days.

Immunocompromised patients are recommended an additional primary series dose (i.e. three doses) compared to immunocompetent patients (i.e. two doses). Thus, vaccination status at index was determined based on immune system status, where patients were classified as immunocompromised at the time of receipt of first COVID-19 vaccine dose if they had one or more codes meeting Davidson *et al*.’s [18] definition of immunocompromised status.

For immunocompetent patients, vaccination status at index (date of COVID-19 diagnosis) was defined according to whether they had received 0 doses (unvaccinated) or 2 primary doses [19]. For immunocompromised patients, vaccination status was defined according to whether they had received 0 doses (unvaccinated) or 3 primary doses [20]. Partially vaccinated patients were excluded.

### Outcomes and follow-up

#### Primary analysis: Long COVID

The primary outcome definition of long COVID was defined as having ≥1 long COVID signs or symptoms as identified by Subramanian et al [3], or a long COVID primary care clinical code (diagnostic or referral code) ≥12 weeks after the initial COVID-19 diagnosis. See **Supplementary file 1** for code lists used. All persons were required to have a minimum of 12 weeks of follow-up. Time at risk commenced 12 weeks after the date of COVID-19 diagnosis until long COVID diagnosis, or persons were censored at the earliest of 9-months follow-up (i.e. post COVID-19 diagnosis), reinfection, post-infection vaccination, date of transfer out of practice, death, or 31 March 2022.

### Secondary analysis: Healthcare resource utilisation

#### Primary care consultations

All-cause primary care consultations, consisting of GP and/or nurse consultations, were reported after a patient was identified as having long COVID until a maximum follow-up of 9-months post-index or censoring. This was defined as a maximum of one visit via telephone or face-to-face consultation per person per day, and any additional visits were considered as data capture errors.

#### Direct healthcare costs

Primary care consultations (including GP and nurse visits) were costed using information by the Personal Social Services Research Unit (PSSRU) [21] (see **Supplementary file 2**) and calculated for all-cause primary care consultations after a patient was identified as having long COVID until a maximum follow-up of 9-months post-index or censoring.

### Covariates

The following sociodemographic characteristics were assessed at index: age (18-49; 50-64; 65-74; 75-84; and ≥85 years), sex (male and female), region of GP practice (North East; North West; Yorkshire and The Humber; East Midlands; West Midlands; East of England; South East Coast; South West; and London), ethnicity (White; Black; Asian; Mixed; and other) and social deprivation (measured using quintiles of the 2019 Index of Multiple Deprivation [IMD] score). Clinical characteristics assessed during the baseline period included: smoking status history (current, former and non-smoker); body mass index (BMI) in the 5 years prior to index (underweight (<18.5 kg/m^2^), normal (18.5-24.9), overweight (25.0-29.9); obese (≥30.0); and unknown); Quan-Charlson Comorbidity Index (CCI) 2005 [22] within two years prior to index (CCI score categories: 0; 1-2; and ≥3); persons at higher risk of severe COVID-19 (high-risk and not at high-risk) defined by the UK’s COVID-19 vaccination prioritisation criteria, the Green Book chapter 14a [23]; frailty as per the electronic frailty index (eFI) [24] (fit and frail, with the latter defined as any level of frailty); influenza vaccination within 12 months prior to index (yes/no); and calendar quarter at index (quarter 1 [1^st^ March 2021-31^st^ May 2021]; quarter 2 [1^st^ June 2021-31^st^ August 2021]; quarter 3 [1^st^ September 2021 – 1^st^ December 2021]). Primary care resource use in the 5 years prior to index was also assessed and defined as two separate variables: pre-pandemic (prior to 1^st^ February 2020) and during pandemic GP practice consultations (1^st^ February and onwards).

### Statistical analysis

Baseline characteristics of patients by vaccination status were summarised using descriptive statistics. The number and percentage of patients with long COVID, the total number of primary care consultations, and primary care consultation costs were reported. Crude long COVID incidence rates per 1,000 person-months were calculated. To minimise confounding, entropy balancing weights were generated [25]. To obtain the average treatment effect (ATE), weights in each of the exposed (vaccinated) and unexposed (unvaccinated) groups were directly calibrated to match the distribution of each covariate in the overall study sample using a set of specified moment conditions including the covariates’ mean, standard deviation, and skewness. Covariates included in the weighting and balancing were sex, GP practice region, ethnicity, IMD score, BMI score, Quan-CCI, high-risk status (the Green Book), frailty and influenza vaccination. Poisson regression was used to estimate unadjusted and adjusted incidence rate ratios, with 95% confidence intervals (CI), for long COVID incidence and to correct for overdispersion robust standard errors were used. This method was also used to assess the association between vaccination status and primary care consultations, with the addition of previous primary care resource use in the entropy balancing. All regression modelling adjusted for age, smoking status, calendar quarter at index. People who had missing data for age, sex, region, social deprivation or smoking status were excluded from the model. The absence of codes for comorbidities in the CCI, high risk definition or eFI was assumed to be the absence of the comorbidity, and not missing data. For BMI, an indicator variable was used for missing value as there were demographic and clinical differences between people with and without BMI captured [17].

The following sensitivity analyses were conducted: 1) accelerated failure time (AFT) models were used due to the potential time-varying nature of long COVID to assess whether patterns differed to the main Poisson regression analysis, 2) using an alternative long COVID definition, where long COVID signs and symptoms or a long COVID clinical code were observed ≥4 weeks after the index date, and 3) primary care consultations were restricted to GP/nurse visits with a long COVID clinical code only.

Outcomes were evaluated by immunocompromised status, and high-risk status in the overall and immunocompetent cohorts. Results for <5 patients were suppressed to comply with CPRD reporting rules, with secondary suppression implemented where relevant. All analyses were conducted in STATA V18.0.

## RESULTS

A total of 111,235 adults with COVID-19 were included in this study; of whom, 67.9% (n=75,522) were unvaccinated and 32.1% (n=35,713) had completed the primary series (**Supplementary figure 2**). Similar distributions were observed in the immunocompetent cohort (unvaccinated group: 66.5% [n=70,145]; primary series group 33.5% [n=35,335]). However, among the 5,755 in the immunocompromised group, 93.4% (n=5,377) were unvaccinated, and 6.6% (n=378) completed the primary vaccine series.

Numerical differences in patient baseline characteristics by vaccination status were observed prior to entropy balancing (**Table 1**). In the overall cohort, and stratified by immune system status, those who completed the primary series were older, more likely White ethnicity, less deprived, less likely in London and more likely in South East than the unvaccinated group. When assessing the sociodemographic and clinical characteristics, patients who had completed the primary series were less likely current smokers, more likely overweight or obese, had greater comorbidity burden, more frail and at higher risk of severe COVID-19. See **Supplementary table 1** for weighted characteristics and **Supplementary table 2** for other unweighted baseline characteristics.

**Table 1.**
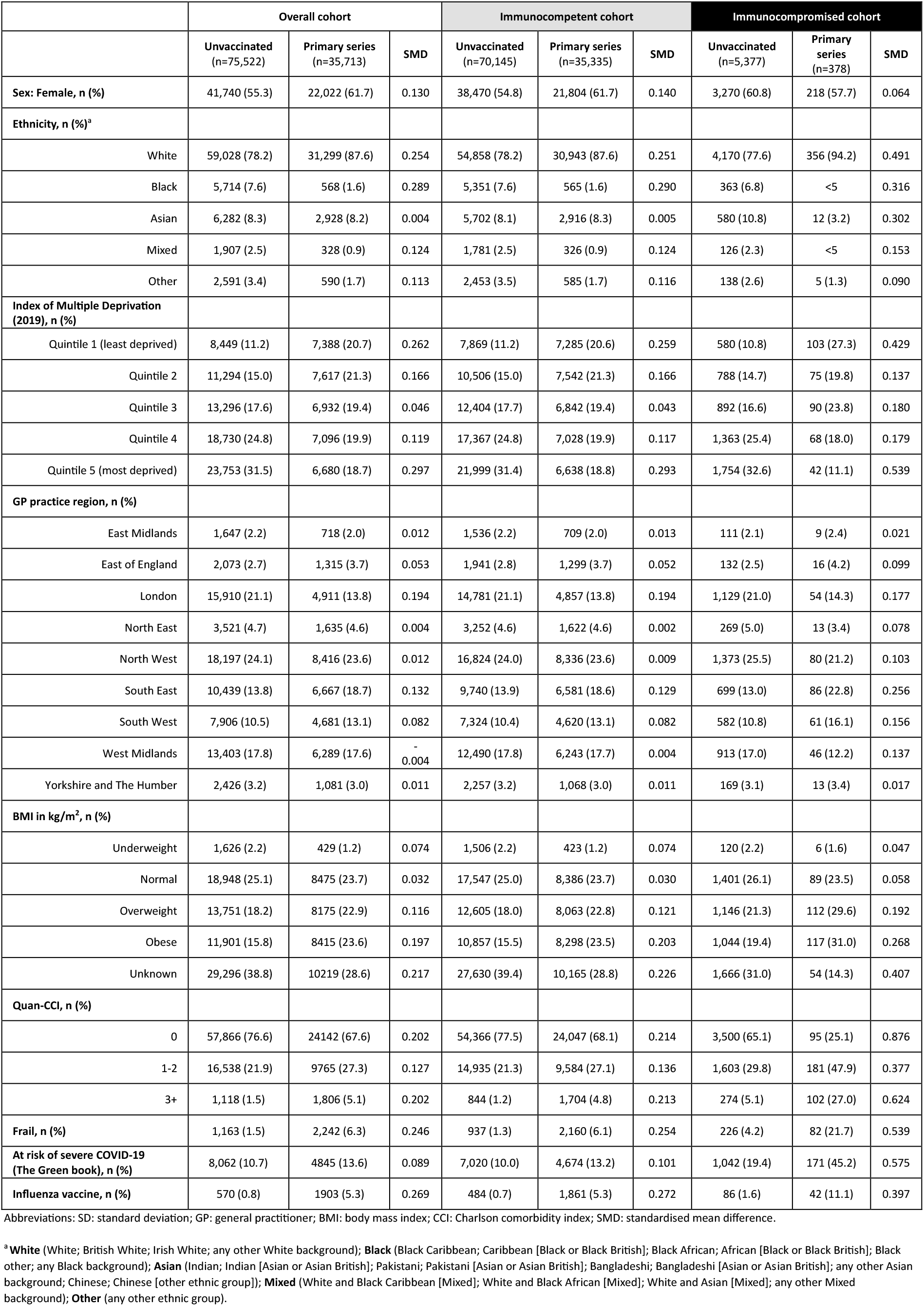
Unweighted sociodemographic and clinical characteristics by vaccination status in the overall, immunocompromised and immunocompetent cohorts.

### Primary analysis

In the overall cohort, the crude long COVID incidence rate per 1,000 person-months was higher for the primary series group than the unvaccinated group, yielding a crude incidence rate ratio (IRR) greater than 1.0 for the primary series group (**Table 2**). However, after entropy balancing and further adjusting for covariates, the incidence rate of long COVID among patients who completed the primary series was significantly lower than unvaccinated patients (adjusted IRR: 0.81; 95% CI: 0.77-0.86).

**Table 2.**
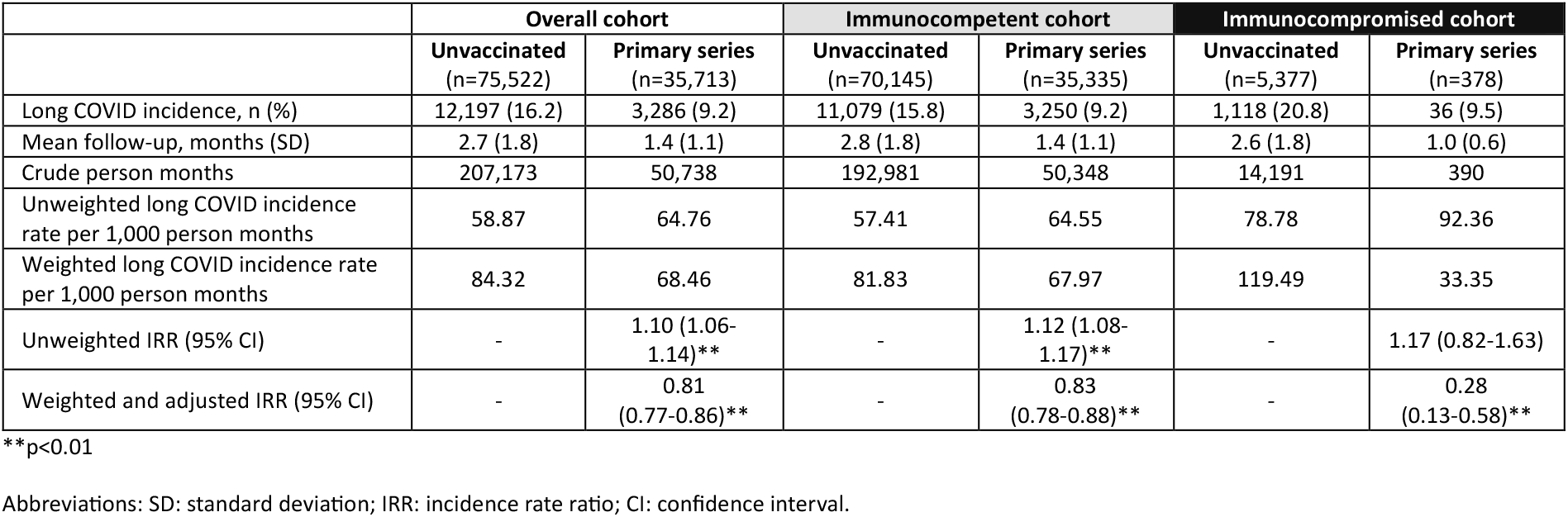
Incidence rates and incidence rate ratio for long COVID, by immune system status.

|  | Overall cohort |  | Immunocompetent cohort |  | Immunocompromised cohort |  |
| --- | --- | --- | --- | --- | --- | --- |
|  | Unvaccinated<br>(n=75,522) | Primary series<br>(n=35,713) | Unvaccinated<br>(n=70,145) | Primary series<br>(n=35,335) | Unvaccinated<br>(n=5,377) | Primary series<br>(n=378) |
| Long COVID incidence, n (%) | 12,197 (16.2) | 3,286 (9.2) | 11,079 (15.8) | 3,250 (9.2) | 1,118 (20.8) | 36 (9.5) |
| Mean follow-up, months (SD) | 2.7 (1.8) | 1.4 (1.1) | 2.8 (1.8) | 1.4 (1.1) | 2.6 (1.8) | 1.0 (0.6) |
| Crude person months | 207,173 | 50,738 | 192,981 | 50,348 | 14,191 | 390 |
| Unweighted long COVID incidence rate per 1,000 person months | 58.87 | 64.76 | 57.41 | 64.55 | 78.78 | 92.36 |
| Weighted long COVID incidence rate per 1,000 person months | 84.32 | 68.46 | 81.83 | 67.97 | 119.49 | 33.35 |
| Unweighted IRR (95% CI) | - | 1.10 (1.06-1.14)** | - | 1.12 (1.08-1.17)** | - | 1.17 (0.82-1.63) |
| Weighted and adjusted IRR (95% CI) | - | 0.81 (0.77-0.86)** | - | 0.83 (0.78-0.88)** | - | 0.28 (0.13-0.58)** |

\*\*p<0.01
Abbreviations: SD: standard deviation; IRR: incidence rate ratio; CI: confidence interval.

#### Long COVID incidence by immune system status

When stratified by immune system status, similar patterns were observed in the immunocompetent cohort: IRR=0.83; 95% CI: 0.78-0.88. Within the immunocompromised cohort a stronger association was noted, as were wide confidence intervals: IRR=0.28; 95% CI: 0.13-0.58 (**Table 2**).

When applying the sensitivity analyses using AFT modelling, to assess the association between long COVID incidence and vaccination status, similar patterns were noted as observed in the main analysis [**Supplementary figures 3** and **4**, and **Supplementary table 3**]. Separately, when using the alternative long COVID definition similar patterns were observed in the overall and immunocompetent cohorts, but no significant association between vaccination status and long COVID was found in the immunocompromised cohort (**Supplementary table 4**).

#### Long COVID incidence by immune system status and high-risk status

Among those at high-risk, weighted and adjusted estimates demonstrated a significant reduction in long COVID IRR with primary series for the overall cohort (IRR=0.82; 95% CI 0.70-0.95) and immunocompetent cohort (IRR=0.82, 95% CI 0.70-0.96) (**Table 3**, immunocompromised cohort not shown due to concordance with high risk definition). Similar patterns were observed among those who were not at high-risk (**Supplementary table 5**).

**Table 3.** Incidence rates and incidence rate ratio for long COVID among those at a higher risk of severe COVID-19 in the overall and immunocompetent cohorts.

|  | Overall high-risk cohort |  | Patients at high-risk within the immunocompetent cohort |  |
| --- | --- | --- | --- | --- |
|  | Unvaccinated<br>(n=8,062) | Primary series<br>(n= 4,845) | Unvaccinated<br>(n= 7,020) | Primary series<br>(n= 4,674) |
| Long COVID incidence, n (%) | 2,036 (25.3) | 661 (13.6) | 1,735 (24.7) | 636 (13.6) |
| Mean follow-up, months (SD) | 2.5 (1.7) | 1.3 (1.0) | 2.5 (1.7) | 1.3 (1.0) |
| Crude person months | 20,131 | 6,300 | 17,553 | 6,138 |
| Unweighted long COVID incidence rate per 1,000 person months | 101.14 | 104.92 | 98.84 | 103.62 |
| Weighted long COVID incidence rate per 1,000 person months | 138.82 | 113.15 | 136.16 | 111.71 |
| Unweighted and unadjusted IRR (95% CI) | - | 1.04<br>(0.95-1.13) | - | 1.05<br>(0.96-1.15) |
| Weighted and adjusted IRR (95% CI) | - | 0.82<br>(0.70-0.95)** | - | 0.82<br>(0.70-0.96)* |

\*p<0.05; \*\*p<0.01
Abbreviations: SD: standard deviation; IRR: incidence rate ratio; CI: confidence interval.

### Secondary analysis

#### The association between vaccination status and primary care consultations in patients with long COVID by immune system status and high-risk status

No significant associations were found between vaccination status and primary care consultation rate in the overall and immunocompetent cohorts (**Table 4**). These findings were also observed when applying the alternative long COVID definition sensitivity analysis (**Supplementary table 6**). When restricting primary care visits to those with a long COVID clinical code only, we observed significantly greater primary care consultations among the primary series group when compared to the unvaccinated group, in the overall (IRR: 1.15; 95% CI: 1.11-1.21) and immunocompetent cohorts (IRR: 1.16; 95% CI: 1.11-1.21) (**Supplementary table 7**). Only descriptive analyses were reported for the immunocompromised cohort due to the small sample size for the primary series group (n=36). Among those at high-risk, no significant association was found between vaccination status and primary care consultation rate in the overall high-risk cohort and immunocompetent cohorts, with similar patterns observed in those who were not at high-risk (**Supplementary table 8**).

**Table 4.**
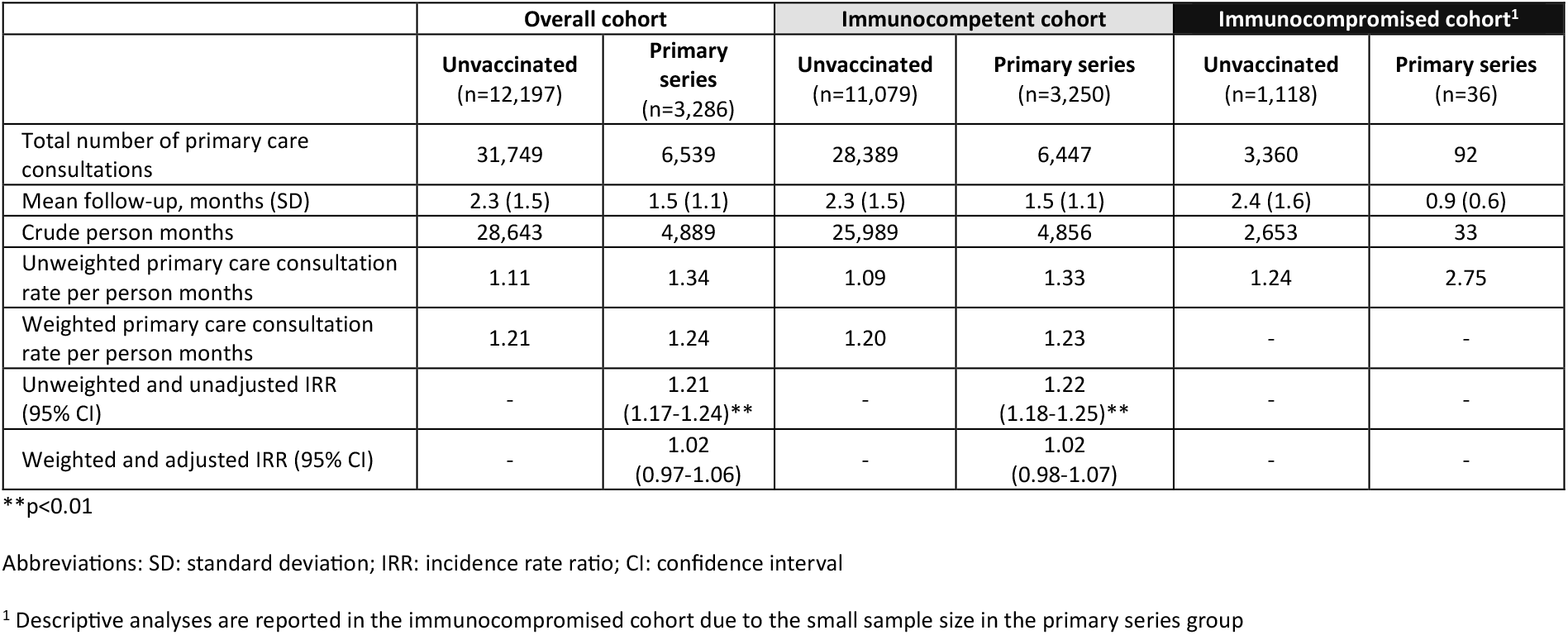
Primary care consultation rates and incidence rate ratio in long COVID patients, by immune system status.

|  | Overall cohort |  | Immunocompetent cohort |  | Immunocompromised cohort<sup>1</sup> |  |
| --- | --- | --- | --- | --- | --- | --- |
|  | Unvaccinated<br>(n=12,197) | Primary series<br>(n=3,286) | Unvaccinated<br>(n=11,079) | Primary series<br>(n=3,250) | Unvaccinated<br>(n=1,118) | Primary series<br>(n=36) |
| Total number of primary care consultations | 31,749 | 6,539 | 28,389 | 6,447 | 3,360 | 92 |
| Mean follow-up, months (SD) | 2.3 (1.5) | 1.5 (1.1) | 2.3 (1.5) | 1.5 (1.1) | 2.4 (1.6) | 0.9 (0.6) |
| Crude person months | 28,643 | 4,889 | 25,989 | 4,856 | 2,653 | 33 |
| Unweighted primary care consultation rate per person months | 1.11 | 1.34 | 1.09 | 1.33 | 1.24 | 2.75 |
| Weighted primary care consultation rate per person months | 1.21 | 1.24 | 1.20 | 1.23 | - | - |
| Unweighted and unadjusted IRR (95% CI) | - | 1.21<br>(1.17-1.24)** | - | 1.22<br>(1.18-1.25)** | - | - |
| Weighted and adjusted IRR (95% CI) | - | 1.02<br>(0.97-1.06) | - | 1.02<br>(0.98-1.07) | - | - |

\*\*p<0.01
Abbreviations: SD: standard deviation; IRR: incidence rate ratio; CI: confidence interval
<sup>1</sup> Descriptive analyses are reported in the immunocompromised cohort due to the small sample size in the primary series group

#### Primary care consultation costs in long COVID patients by immune system status and high-risk status

The overall mean costs were higher in the unvaccinated group than the group who had received the primary series (**Table 5**). These costs were similar in the immunocompetent cohort but were more pronounced in the immunocompromised cohort. Comparable patterns were observed when stratified by high-risk status, and separately, when applying the sensitivity analyses by restricting primary care visits to those with a long COVID clinical code only (**Supplementary table 9**). However, patterns were less clear when applying the alternative long COVID definition (**Supplementary table 10**).

**Table 5.** Primary care consultation costs in long COVID patients by immune system status and high-risk status.

|  | Overall cohort |  | Immunocompetent cohort |  | Immunocompromised cohort |  |
| --- | --- | --- | --- | --- | --- | --- |
|  | Unvaccinated<br>(n=12,197) | Primary series<br>(n=3,286) | Unvaccinated<br>(n=11,079) | Primary series<br>(n=3,250) | Unvaccinated<br>(n=1,118) | Primary series<br>(n=36) |
| Overall |  |  |  |  |  |  |
| Mean primary care consultation costs, £ (SD) | 55.6 (59.7) | 43.1 (45.5) | 54.7 (59.2) | 43.0 (45.3) | 64.2 (64.2) | 53.6 (62.1) |
| Median primary care consultation costs, £ (Q1, Q3) | 39.2 (15.5, 75.6) | 31.0 (15.5, 54.8) | 39.2 (15.5, 70.3) | 31.0 (15.5, 54.8) | 44.6 (15.5, 85.8) | 31.0 (15.5, 66.4) |
| Patients at a higher risk of severe COVID-19 |  |  |  |  |  |  |
|  | Unvaccinated<br>(n=2,036) | Primary series<br>(n=661) | Unvaccinated<br>(n=1,735) | Primary series<br>(n=636) | n/a |  |
| Mean primary care consultation costs, £ (SD) | 70.1 (78.2) | 49.5 (56.8) | 69.5 (79.5) | 48.9 (56.2) |  |  |
| Median primary care consultation costs, £ (Q1, Q3) | 46.6 (15.5, 93.7) | 39.2 (15.5, 62.1) | 46.6 (15.5, 92.6) | 39.2 (15.5, 62.1) |  |  |
| Patients not at a higher risk of severe COVID-19 |  |  |  |  |  |  |
|  | Unvaccinated<br>(n=10,161) | Primary series<br>(n=2,625) | Unvaccinated<br>(n=9,344) | Primary series<br>(n=2,614) | n/a |  |
| Mean primary care consultation costs, £ (SD) | 52.6 (54.8) | 41.5 (42.1) | 51.9 (54.1) | 41.6 (42.1) |  |  |
| Median primary care consultation costs, £ (Q1, Q3) | 39.2 (15.5, 70.3) | 31.0 (15.5, 54.8) | 39.2 (15.5, 70.3) | 31.0 (15.5, 54.8) |  |  |

Abbreviations: £: British pound sterling; SD: standard deviation; Q1: quartile 1; Q3: quartile 3; n/a: not applicable.

## DISCUSSION

In this study, completion of a COVID-19 vaccine primary series was associated with a lower incidence rate of long COVID. The association was more pronounced, although with wide confidence intervals, in the immunocompromised cohort. These findings were unchanged when assessed among those at high-risk of severe COVID-19. Among those with long COVID, there was no difference in primary care consultations between vaccinated and unvaccinated persons.

Our findings align with a recent systematic review [26], observational evidence from three European countries [27] and observational evidence with a focus on specific post-COVID-19 conditions [28, 29] on the potential preventative effects of primary series vaccination on developing long COVID. However, to our knowledge this is the first study to assess the effects of vaccination on long COVID related HCRU and costs by immune system status. While our findings showed a stronger positive association of primary series vaccination in immunocompromised patients than in the overall cohort it should be noted that a relatively small (n=378; 6.6%) group of immunocompromised patients had received the primary series at index, which was proportionally a much lower number of vaccinated patients than in the overall and immunocompetent cohorts. When applying the alternative long COVID definition (signs/symptoms or long COVID diagnosis code at ≥4 weeks post index) no evidence of an association was found in the immunocompromised cohort. Nevertheless, evidence on the risk of developing long COVID by immune system status is limited [30], and our study supports a potential protective effect of COVID-19 vaccination against long COVID in both immunocompromised and immunocompetent patients. The reduced long COVID IRRs observed in immunocompromised patients may in-part be explained by the intrinsic differences between immunocompromised and immunocompetent patients. Immunocompromised patients have an increased susceptibility to developing complications from COVID-19, with some patients being more likely to experience persistent SARS-CoV-2 infections [31-33]; this may translate into severe long COVID presentations in certain immunocompromised patients.

The association between vaccination status and long COVID incidence was not substantially different between those at high risk and those who were not at high risk of severe acute COVID. These findings may be attributed to our defined population; we only capture patients in the primary care setting, who were more likely to be mild-to-moderate long COVID cases than those receiving care for COVID-19 in hospital, whilst a stronger vaccination impact has been previously observed in severe long COVID cases with functional impairment [34].

No association was observed between primary series vaccination and rate of all-cause primary care consultations in the overall and immunocompetent cohorts. When the definition of primary care consultations was limited to people with a long COVID clinical code, we observed a significantly greater rate of resource use in the primary series group compared to the unvaccinated group in both the overall and immunocompetent cohorts. Recent evidence in other therapy areas has shown a strong correlation between frequency of GP visits and vaccination uptake [35]; hence greater frequency of long COVID related GP visits in the vaccinated group may reflect greater health seeking behaviour rather than greater resource burden resulting from long COVID, especially when considering that our definition of long COVID is dependent on patients reporting signs and symptoms in primary care.

Direct healthcare costs were numerically greater in unvaccinated adults than among those who had received the primary series. However, this study evaluated the effect of vaccination on utilisation of primary care services but did not compare the associated primary care costs. Further research is needed to explore whether COVID-19 vaccination reduces long COVID-related costs.

Our study had several limitations. Firstly, the operationalised definition of long COVID applied in this study was based on clinical coded data of signs/symptoms or long COVID diagnosis codes, and therefore may underrepresent the true burden of long COVID. Further, due to the nature of electronic health records we were unable to apply part of WHO’s definition of long COVID: patients with long COVID signs/symptoms persisting for ≥2 months [2] which cannot be explained by alternative diagnosis, and therefore may have misclassified some patients as having long COVID. Evidence using UK primary care records demonstrates only 20% of symptoms are captured as a clinical code, with the remaining 80% as free text in COVID-19 patients [36], which may lead to potential misclassification of patients. As some SARS-CoV-2 cases are missing from primary care records this study may have been affected by selection bias [37], however we would expect this bias to equally affect the primary series and unvaccinated groups, and therefore should have minimal impact on the associations observed. The relatively short indexing period of 9 months may not fully represent patients with diagnosed with COVID-19 during earlier or later time periods. Further, the short follow-up period across both groups, but particularly the primary series group, may mean the full impact of long COVID (both incidence and primary care resource use) may have been underestimated. Prior to entropy balancing, those who had received the primary series were older, more commonly overweight/obese, had greater comorbidity burden, a greater proportion of patients with frailty and at higher-risk of severe COVID-19; such characteristics are associated with greater morbidity and therefore greater need for more frequent healthcare services interaction, which is supported by the higher baseline primary care consultation rates observed in the vaccinated than the unvaccinated group. Where patients more regularly interact with the healthcare system for non-long COVID related reasons, we expect an increased degree of misclassification of diagnoses as long COVID cases, leading to an overestimation of the incidence of long COVID within this group.

Whilst stratification by immune system status is a strength of this study, the sample size of the immunocompromised cohort was small; this particularly affected results for primary care consultations and sensitivity analyses, leading to less precise estimates. To limit confounding, we applied entropy balancing and accounted for many important covariates in the regression models. However, residual confounding may nonetheless be present.

## CONCLUSIONS

Our study highlights the importance of continuing efforts with the COVID-19 vaccination in the UK to prevent or reduce the risk of developing long COVID, which may help to alleviate the long COVID economic burden in the primary care setting. It is one of the few studies using national data to characterise long covid by immune status and high-risk status. While the study period was short due to data availability, our novel evidence provides a reference point that was previously lacking, from which further research on this topic with longer follow up can compare against.

## LIST OF ABBREVIATIONS

AFT: accelerated failure time; ATE: average treatment effect; BMI: body mass index; CI: confidence intervals; COVID-19: coronavirus disease 2019; CPRD: Clinical Practice Research Datalink; eFI: electronic frailty index; GP: general practitioner; HES: Hospital Episode Statistics; HES APC: Hospital Episode Statistics Admitted Patient Care; IRR: incidence rate ratio; IMD: Index of Multiple Deprivation; n/a: not applicable; PSSRU: Personal Social Services Research Unit; Quan-CCI: Quan-Charlson Comorbidity Index; Q1: quartile 1; Q3: quartile 3; SARS-CoV-2: Severe acute respiratory syndrome coronavirus 2; SD: standard deviation; SMD: standardised mean difference; UK: United Kingdom: WHO: World Health Organization; £: British pound sterling.

## DECLARATIONS

### Ethics approval and consent to participate

This study involves human participants and was approved by Clinical Practice Research Datalink’s Research Data Governance (RDG): CPRD study ID: 22_002431. This study involved the use of secondary data; no primary data collection was carried out for the purpose of this study. As all patient-level data were fully anonymised, and no direct patient contact or primary collection of individual patient data occurred, patient consent was not required.

### Consent for publication

Not applicable.

## Availability of data and materials

Access to anonymised patient data from CPRD is subject to a data-sharing agreement containing detailed terms and conditions of use following protocol approval from CPRD’s RDG. All data relevant to the study are included in the article or uploaded as supplementary information. Further queries can be directed to the corresponding author at.

## Data Availability

All data produced in the present study are available upon reasonable request to the authors.

## Author contributions

JY conceptualized, designed, interpreted the data, revised and reviewed the manuscript. KKR designed, interpreted the data and drafted the initial manuscript. TA interpreted the data and reviewed the manuscript. LM analysed and interpreted the data, and reviewed the manuscript. OM analysed and interpreted the data, and reviewed the manuscript. LM interpreted the data and reviewed the manuscript. KMA interpreted the data and reviewed the manuscript. TT interpreted the data, revised and reviewed the manuscript. CT interpreted the data and reviewed the manuscript. RB interpreted the data and reviewed the manuscript. CR interpreted the data and reviewed the manuscript. DM interpreted the data and reviewed the manuscript. JLN conceptualized, revised and reviewed the manuscript.

