## Supplementary material for "The impact of COVID vaccination on incidence of long COVID and healthcare resource utilisation in a primary care cohort in England, 2021-2022": Impact of vaccines on long COVID Supp materials V1.0 24042024 .pdf

Supplementary figure 1 Study design schematic

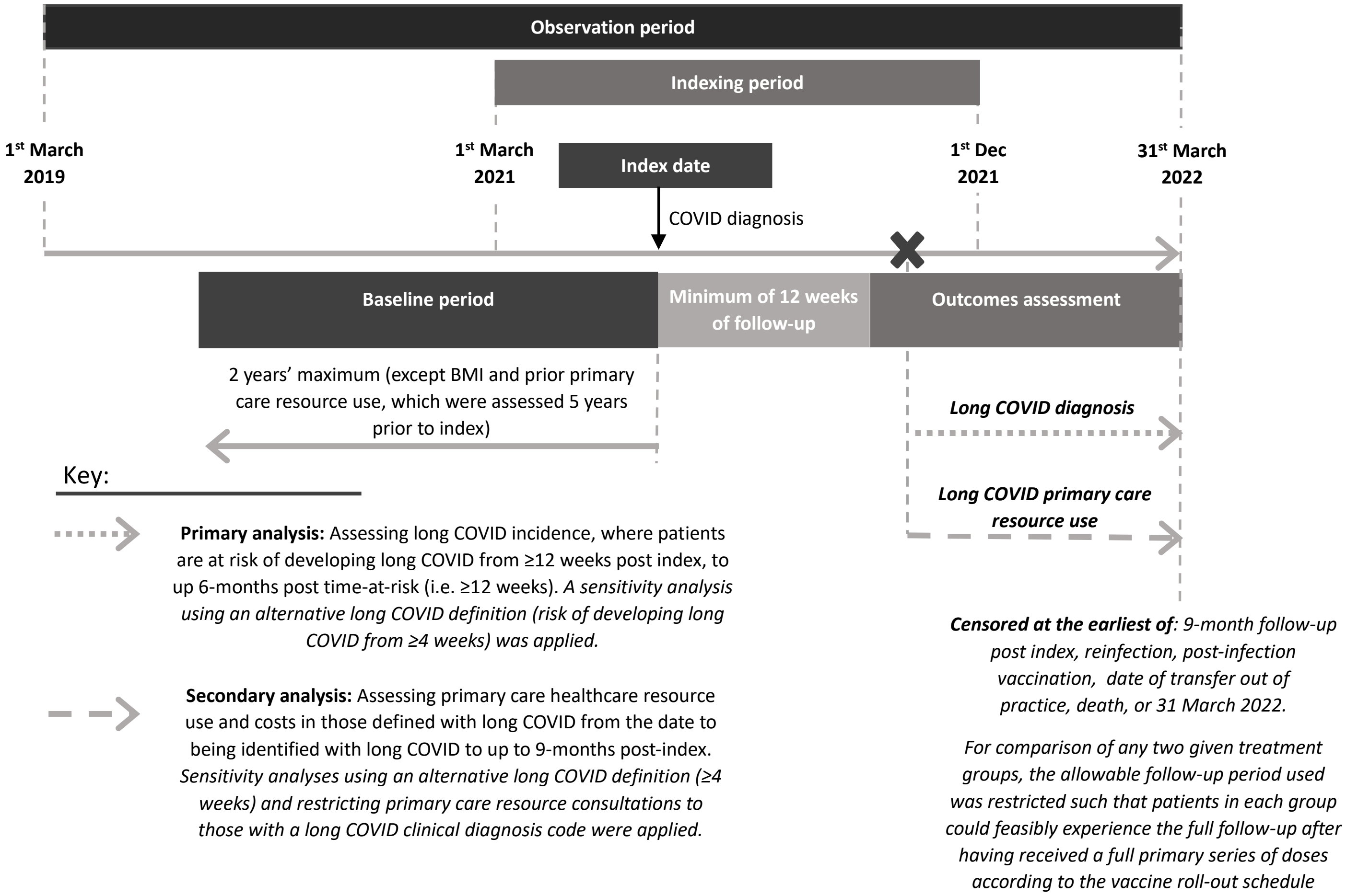

Supplementary figure 2 Patient attrition flow diagram

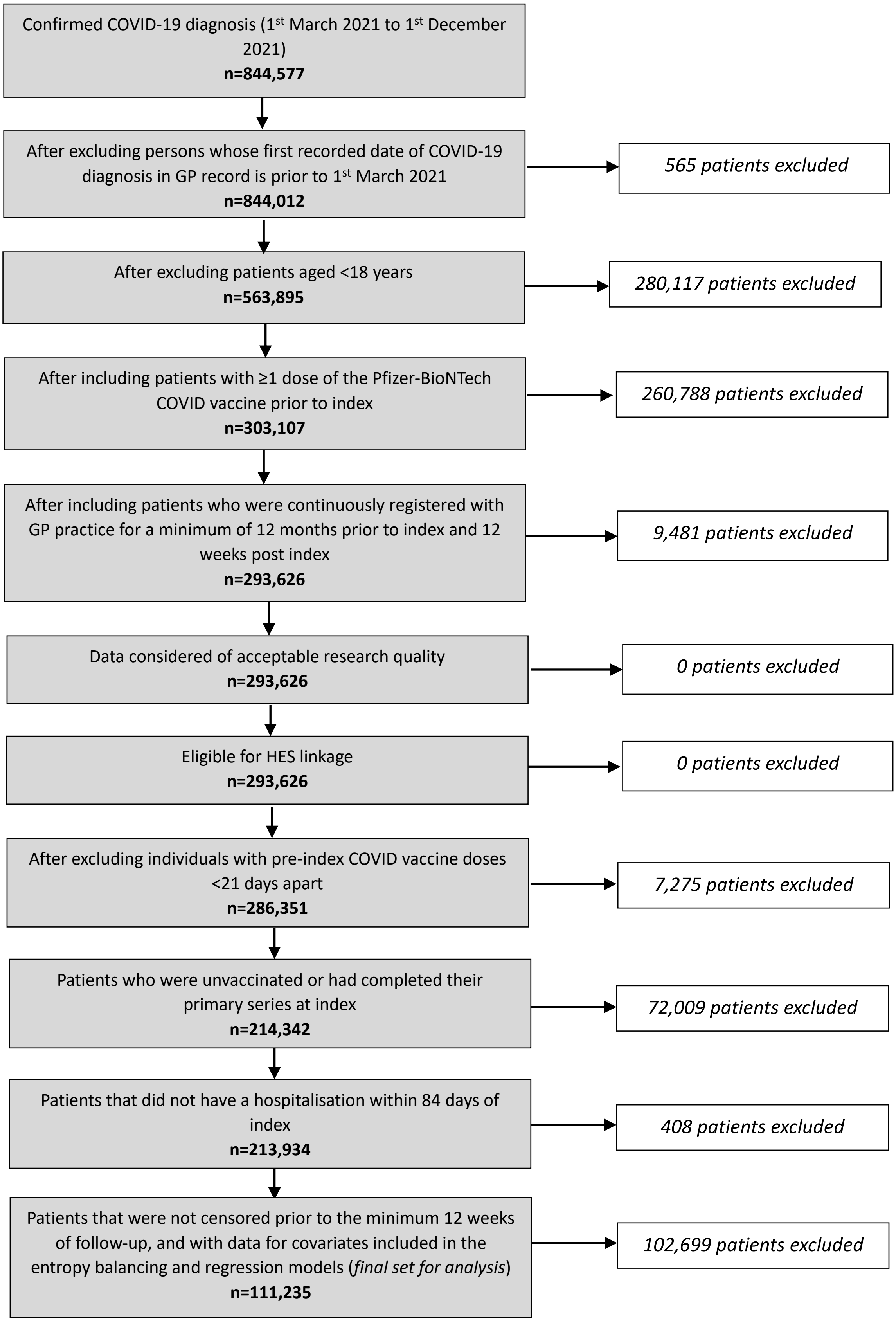

**Supplementary figure 3** Unadjusted (3a) and adjusted (3b) Kaplan-Meier curves from accelerated failure time (AFT) models showing the incidence of long COVID, by vaccination status, in the overall cohort

**Supplementary figure 3a:** Unadjusted Kaplan-Meier curves

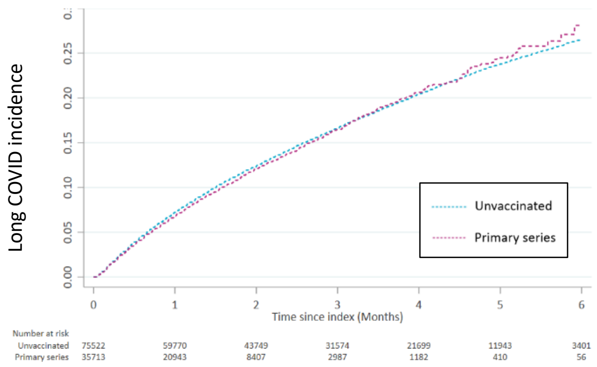

**Supplementary figure 3b:** Adjusted Kaplan-Meier curves

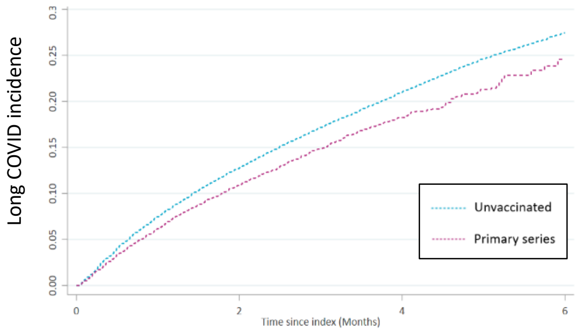

**Supplementary figure 4** Unadjusted (4a) and adjusted (4b) Kaplan-Meier curves showing the incidence of long COVID, by vaccination status, in the immunocompetent cohort

**Supplementary figure 4a:** Unadjusted Kaplan-Meier curves

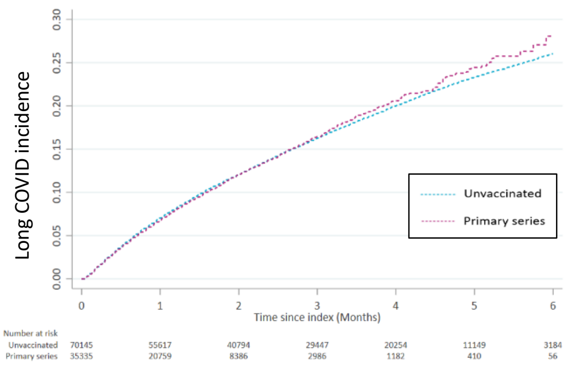

**Supplementary figure 4b:** Adjusted Kaplan-Meier curves

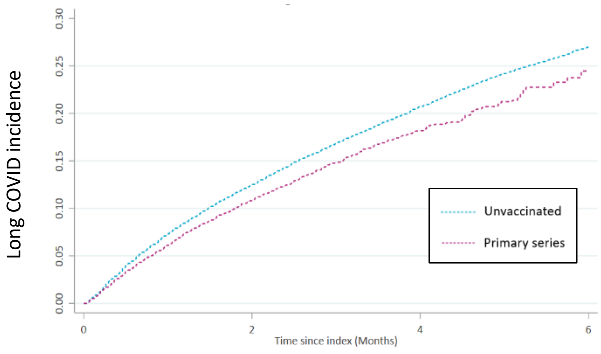

**Supplementary table 1** Weighted sociodemographic and clinical characteristics by vaccination status in the overall, immunocompromised and immunocompetent cohorts for the primary analysis

| | Overall cohort | | | Immunocompetent cohort | | | Immunocompromised cohort | | |
|---|---|---|---|---|---|---|---|---|---|
| | **Unvaccinated** (n=75,522) | **Primary series** (n=35,713) | | **Unvaccinated** (n=70,145) | **Primary series** (n=35,335) | | **Unvaccinated** (n=5,377) | **Primary series** (n=378) | |
| **Sex: female, n (%)** | 41,740 (57.3) | 22022 (57.3) | <0.001 | 38,470 (57.1) | 21,804 (57.1) | <0.001 | 3,270 (60.6) | 218 (60.6) | <0.001 |
| **Ethnicity, n (%)[a]** | | | | | | | | | |
| White | 59,028 (81.2) | 3,1299 (81.2) | <0.001 | 54,858 (81.3) | 30,943 (81.3) | <0.001 | 4,170 (78.6) | 356 (78.6) | 0 |
| Black | 5,714 (5.7) | 568 (5.7) | <0.001 | 5,351 (5.6) | 565 (5.6) | <0.001 | 363 (6.4) | 3 (6.4) | <0.001 |
| Asian | 6,282 (8.3) | 2,928 (8.8) | <0.001 | 5,702 (8.2) | 2,916 (8.2) | <0.001 | 580 (10.3) | 12 (10.3) | <0.001 |
| Mixed | 1,907 (2.0) | 328 (2.0) | <0.001 | 1,781 (2.0) | 326 (2.0) | <0.001 | 126 (2.2) | 2 (2.2) | <0.001 |
| Other | 2,591 (2.9) | 590 (2.9) | <0.001 | 2,453 (2.9) | 585 (2.9) | <0.001 | 138 (2.5) | 5 (2.5) | <0.001 |
| **Index of Multiple Deprivation (2019), n (%)** | | | | | | | | | |
| Quintile 1 (least deprived) | 8,449 (14.2) | 7,388 (14.2) | <0.001 | 7,869 (14.4) | 7,285 (14.4) | <0.001 | 580 (11.9) | 103 (11.9) | <0.001 |
| Quintile 2 | 11,294 (17.0) | 7,617 (17.0) | <0.001 | 10,506 (17.1) | 7,542 (17.1) | <0.001 | 788 (15.0) | 75 (15.0) | 0 |
| Quintile 3 | 13,296 (18.2) | 6,932 (18.2) | <0.001 | 12,404 (18.3) | 6,842 (18.3) | <0.001 | 892 (17.1) | 90 (17.1) | <0.001 |
| Quintile 4 | 18,730 (23.2) | 7,096 (23.2) | <0.001 | 17,367 (23.1) | 7,028 (23.1) | <0.001 | 1,363 (24.9) | 68 (24.9) | <0.001 |
| Quintile 5 (most deprived) | 23,753 (27.4) | 6,680 (27.4) | <0.001 | 21,999 (27.2) | 6,638 (27.2) | <0.001 | 1,754 (31.2) | 42 (31.2) | <0.001 |
| **GP practice region, n (%)** | | | | | | | | | |
| East Midlands | 1,647 (2.1) | 718 (2.1) | <0.001 | 1,536 (2.1) | 709 (2.1) | <0.001 | 111 (2.1) | 9 (2.1) | <0.001 |
| East of England | 2,073 (3.1) | 1,315 (3.1) | <0.001 | 1,941 (3.1) | 1,299 (3.1) | <0.001 | 132 (2.6) | 16 (2.6) | <0.001 |
| London | 15,910 (18.7) | 4,911 (18.7) | <0.001 | 14,781 (18.6) | 4,857 (18.6) | <0.001 | 1,129 (20.6) | 54 (20.6) | <0.001 |
| North East | 3,521 (4.6) | 1,635 (4.6) | <0.001 | 3,252 (4.6) | 1,622 (4.6) | <0.001 | 269 (4.9) | 13 (4.9) | 0 |
| North West | 18,197 (23.9) | 8,416 (23.9) | <0.001 | 16,824 (23.9) | 8,336 (23.9) | <0.001 | 1,373 (25.3) | 80 (25.3) | <0.001 |
| South East | 10,439 (15.4) | 6,667 (15.4) | <0.001 | 9,740 (15.5) | 6,581 (15.5) | <0.001 | 699 (13.6) | 86 (13.6) | <0.001 |
| South West | 7,906 (11.3) | 4,681 (11.3) | <0.001 | 7,324 (11.3) | 4,620 (11.3) | <0.001 | 582 (11.2) | 61 (11.2) | 0 |
| West Midlands | 13,403 (17.7) | 6,289 (17.7) | <0.001 | 12,490 (17.8) | 6,243 (17.8) | <0.001 | 913 (16.7) | 46 (16.7) | <0.001 |
| Yorkshire and The Humber | 2,426 (3.2) | 1,081 (3.2) | <0.001 | 2,257 (3.2) | 1,068 (3.2) | <0.001 | 169 (3.2) | 13 (3.2) | <0.001 |
| **BMI in kg/m$^2$, n (%)** | | | | | | | | | |
| Underweight | 1,626 (1.9) | 429 (1.9) | <0.001 | 1,506 (1.8) | 423 (1.8) | <0.001 | 120 (2.2) | 6 (2.2) | <0.001 |
| Normal | 18,948 (24.7) | 8,475 (24.7) | <0.001 | 17,547 (24.6) | 8,386 (24.6) | <0.001 | 1,401 (25.9) | 89 (25.9) | <0.001 |
| Overweight | 13,751 (19.7) | 8,175 (19.7) | <0.001 | 12,605 (19.6) | 8,063 (19.6) | <0.001 | 1,146 (21.9) | 112 (21.9) | 0 |
| Obese | 11,901 (18.3) | 8,415 (18.3) | <0.001 | 10,857 (18.2) | 8,298 (18.2) | <0.001 | 1,044 (20.2) | 117 (20.2) | <0.001 |
| Unknown | 29,296 (35.5) | 10,219 (35.5) | <0.001 | 27,630 (35.8) | 10,165 (35.8) | <0.001 | 1,666 (29.9) | 54 (29.9) | <0.001 |
| **Quan-CCI, n (%)** | | | | | | | | | |
| 0 | 57,866 (73.7) | 24,142 (73.7) | 0 | 54,366 (74.3) | 24,047 (74.3) | <0.001 | 3,500 (62.5) | 95 (62.5) | <0.001 |
| 1-2 | 16,538 (23.7) | 9,765 (23.7) | <0.001 | 14,935 (23.3) | 9,584 (23.3) | <0.001 | 1,603 (31.0) | 181 (31.0) | 0 |
| 3+ | 1,118 (2.6) | 1,806 (2.6) | <0.001 | 844 (2.4) | 1,704 (2.4) | <0.001 | 274 (6.5) | 102 (6.5) | <0.001 |
| **Frail, n (%)** | 1,163 (3.1) | 2,242 (3.1) | <0.001 | 937 (2.9) | 2,160 (2.9) | <0.001 | 226 (5.4) | 82 (5.4) | 0 |
| **At risk of severe COVID-19 (The Green book), n (%)** | 8,062 (11.6) | 4,845 (11.6) | <0.001 | 7,020 (11.1) | 4,674 (11.1) | <0.001 | 1,042 (21.1) | 171 (21.1) | <0.001 |
| **Influenza vaccine, n (%)** | 570 (2.2) | 1,903 (2.2) | <0.001 | 484 (2.2) | 1,861 (2.2) | <0.001 | 86 (2.2) | 42 (2.2) | 0 |

Abbreviations: SD: standard deviation; GP: general practitioner; BMI: body mass index; CCI: Charlson comorbidity index; SMD: standardised mean difference.

[a] **White** (White; British White; Irish White; any other White background); **Black** (Black Caribbean; Caribbean [Black or Black British]; Black African; African [Black or Black British]; Black other; any Black background); **Asian** (Indian; Indian [Asian or Asian British]; Pakistani; Pakistani [Asian or Asian British]; Bangladeshi; Bangladeshi [Asian or Asian British]; any other Asian background; Chinese; Chinese [other ethnic group]); **Mixed** (White and Black Caribbean [Mixed]; White and Black African [Mixed]; White and Asian [Mixed]; any other Mixed background); **Other** (any other ethnic group).

Note: The following covariates did not feature in the entropy balancing and were adjusted for in the regression models: age, smoking status and index quarter.

Supplementary table 2: Additional information on unweighted sociodemographic and clinical characteristics by vaccination status in the overall, immunocompromised and immunocompetent cohorts for the primary analysis

| | Overall cohort | | Immunocompetent cohort | | Immunocompromised cohort | |
|---|---|---|---|---|---|---|
| | **Unvaccinated** (n=75,522) | **Primary series** (n=35,713) | **Unvaccinated** (n=70,145) | **Primary series** (n=35,335) | **Unvaccinated** (n=5,377) | **Primary series** (n=378) |
| **Age at index in years, mean (SD)** | 34.4 (11.4) | 44.6 (15.8) | 34.1 (11.2) | 44.4 (15.8) | 38.2 (12.7) | 57.8 (15.6) |
| **Age at index in years, n (%)** | | | | | | |
| 18-49 | 67,492 (89.4) | 23,759 (66.5) | 63,049 (89.9) | 23,633 (66.9) | 4,443 (82.6) | 126 (33.3) |
| 50-64 | 6,832 (9.1) | 7,166 (20.1) | 6,097 (8.7) | 7,046 (19.9) | 735 (13.7) | 120 (31.8) |
| 65-74 | 829 (1.1) | 2,899 (8.1) | 706 (1.0) | 2,828 (8.0) | 123 (2.3) | 71 (18.8) |
| 75-84 | 259 (0.3) | 1,492 (4.2) | 205 (0.3) | 1,442 (4.1) | 54 (1.0) | 50 (13.2) |
| ≥85 | 110 (0.2) | 397 (1.1) | 88 (0.1) | 386 (1.1) | 22 (0.4) | 11 (2.9) |
| **Smoking status, n (%)** | | | | | | |
| Current smoker | 24,746 (32.8) | 7376 (20.7) | 22,885 (32.6) | 7,315 (20.7) | 1,861 (34.6) | 61 (16.1) |
| Former smoker | 21,906 (29.0) | 14389 (40.3) | 20,020 (28.5) | 14,180 (40.1) | 1,886 (35.1) | 209 (55.3) |
| Non-smoker | 28,870 (38.2) | 13948 (39.1) | 27,240 (38.8) | 13,840 (39.2) | 1,630 (30.3) | 108 (28.6) |
| **BMI in kg/m$^2$, mean (SD)** | 26.9 (6.4) | 28.4 (6.7) | 26.9 (6.4) | 28.4 (6.7) | 27.4 (6.6) | 28.6 (6.0) |
| **Quan-CCI, mean (SD)** | 0.3 (0.8) | 0.6 (1.2) | 0.3 (0.7) | 0.6 (1.2) | 0.6 (1.3) | 2.0 (2.3) |
| **Index quarter, n (%)[a]** | | | | | | |
| Quarter 1 | 6894 ( 9.1) | 278 ( 0.8) | 6390 ( 9.1) | 278 ( 0.8) | 504 ( 9.4) | <5 |
| Quarter 2 | 39165 (51.9) | 12589 (35.3) | 36506 (52.0) | 12588 (35.6) | 2659 (49.5) | <5 |
| Quarter 3 | 29463 (39.0) | 22846 (64.0) | 27249 (38.8) | 22469 (63.6) | 2214 (41.2) | 377 (99.7) |
| **GP practice consultations per person-months, pre-pandemic, n (%)** | | | | | | |
| ≤0.1 | 18,777 (24.9) | 5,812 (16.3) | 18,058 (25.7) | 5,796 (16.4) | 719 (13.4) | 16 ( 4.2) |
| >0.1-0.3 | 23,766 (31.5) | 10,965 (30.7) | 22,270 (31.7) | 10,884 (30.8) | 1,496 (27.8) | 81 (21.4) |
| >0.3-≤0.5 | 13,635 (18.1) | 7,755 (21.7) | 12,484 (17.8) | 7,687 (21.8) | 1,151 (21.4) | 68 (18.0) |
| >0.5 | 16,457 (21.8) | 9,809 (27.5) | 14,620 (20.8 | 9,610 (27.2) | 1,837 (34.2) | 199 (52.6) |
| Missing | 2,887 (3.8) | 1,372 (3.8) | 2,713 ( 3.9) | 1,358 ( 3.8) | 174 ( 3.2) | 14 ( 3.7) |
| **GP practice consultations per person-months during the pandemic, n (%)** | | | | | | |
| ≤0.1 | 26,976 (35.7) | 9,377 (26.3) | 25,762 (36.7) | 9,340 (26.4) | 1,214 (22.6) | 37 ( 9.8) |
| >0.1-0.2 | 12,937 (17.1) | 6,086 (17.0) | 12,116 (17.3) | 6,055 (17.1) | 821 (15.3) | 31 ( 8.2) |
| >0.2-≤0.4 | 15,399 (20.4) | 8,601 (24.1) | 14,207 (20.3) | 8,500 (24.1) | 1,192 (22.2) | 101 (26.7) |
| >0.4 | 20,210 (26.8) | 11,649 (32.6) | 18,060 (25.7) | 11,440 (32.4) | 2,150 (40.0) | 209 (55.3) |

Abbreviations: SD: standard deviation; GP: general practitioner; BMI: body mass index; CCI: Charlson comorbidity index

[a] Index quarters were categorised as follows: **quarter 1**: 1st March 2021-31st May 2021; **quarter 2**: 1st June 2021- 31st August 2021; and **quarter 3**: 1st September 2021-1st December 2021.

**Supplementary table 3:** Time ratio estimates from accelerated failure time (AFT) models for time to developing long COVID: comparing the unvaccinated group (reference) to those who completed primary series vaccination

| | Ratio of time to long COVID between those with the primary series and those who are unvaccinated | | |
|---|---|---|---|
| | **Overall cohort** | **Immunocompetent cohort** | **Immunocompromised cohort** |
| Unweighted time ratio (95% CI) | 0.96<br>(0.92-1.00) | 0.94<br>(0.90-0.98)** | NR |
| Weighted and adjusted time ratio (95% CI) | 1.17<br>(1.12-1.22)** | 1.14<br>(1.09-1.20)** | NR |

Note: A coefficient (time ratio) >1.0 indicates the effect of the exposure increases survival time (i.e. a longer duration in developing long COVID. Whereas a coefficient <1.0 indicates decreases in time to attrition (i.e. speeds up the time for developing long COVID). AFT modelling results were uninterpretable and therefore not reported in the immunocompromised cohort due to the limited sample size.

**p<0.01

Abbreviations: CI: confidence interval; NR: not reported.

**Supplementary table 4:** Incidence rates and incidence rate ratio for long COVID, by immune system status using an alternative definition of long COVID (signs and symptoms at ≥ 4 weeks)

| | **Overall cohort** | | **Immunocompetent cohort** | | **Immunocompromised cohort** | |
|---|---|---|---|---|---|---|
| | **Unvaccinated** (n=100,874) | **Primary series** (n=63,066) | **Unvaccinated** (n=94,140) | **Primary series** (n=62,615) | **Unvaccinated** (n=6,734) | **Primary series** (n=451) |
| Long COVID incidence, n (%) | 18,526 (18.4) | 8,064 (12.8) | 16,864 (17.9) | 7,972 (12.7) | 1,662 (24.7) | 92 (20.4) |
| Mean follow-up, months (SD) | 3.3 (2.3) | 2.1 (1.4) | 3.3 (2.3) | 2.1 (1.4) | 3.3 (2.3) | 2.2 (1.1) |
| Crude person months | 337,283 | 130,867 | 314,933 | 129,865 | 22,350 | 1,002 |
| Unweighted long COVID incidence rate per 1,000 person months | 54.92 | 61.62 | 53.55 | 61.39 | 74.36 | 91.77 |
| Weighted long COVID incidence rate per 1,000 person months | 81.52 | 66.23 | 80.09 | 66.18 | 106.84 | 69.52 |
| Unweighted and unadjusted IRR (95% CI) | - | 1.12 (1.09-1.15)** | - | 1.15 (1.12-1.18)** | - | 1.23 (0.99-1.52) |
| Weighted and adjusted IRR (95% CI) | - | 0.81 (0.78-0.85) ** | - | 0.83 (0.79-0.86)** | - | 0.65 (0.34-1.23) |

**p<0.01

Abbreviations: SD: standard deviation; IRR: incidence rate ratio; CI: confidence interval.

Supplementary table 5: Incidence rates and incidence rate ratio for long COVID among those not at higher risk of severe COVID-19 in the overall and immunocompetent cohorts

| | **Overall high-risk cohort** | | **Patients not at high-risk within the immunocompetent cohort** | |
|---|---|---|---|---|
| | Unvaccinated (n=67,460) | Primary series (n= 30,868) | Unvaccinated (n=63,125) | Primary series (n=30,661) |
| Long COVID incidence, n (%) | 10,161 (15.1) | 2,625 (8.5) | 9,344 (14.8) | 2,614 (8.5) |
| Mean follow-up, months (SD) | 2.8 (1.8) | 1.4 (1.1) | 2.8 (1.8) | 1.4 (1.1) |
| Crude person months | 187,042 | 44,438 | 175,429 | 44,210 |
| Unweighted long COVID incidence rate per 1,000 person months | 54.32 | 59.07 | 53.26 | 59.13 |
| Weighted long COVID incidence rate per 1,000 person months | 76.61 | 63.10 | 74.16 | 62.84 |
| Unweighted and unadjusted IRR (95% CI) | - | 1.09 (1.04-1.14)** | - | 1.11 (1.06-1.16)** |
| Weighted and adjusted IRR (95% CI) | - | 0.82 (0.77-0.88)** | - | 0.85 (0.80-0.90)** |

**p<0.01

Abbreviations: SD: standard deviation; IRR: incidence rate ratio; CI: confidence interval.

Supplementary table 6: Primary care consultation rates and incidence rate ratio in long COVID patients, by immune system status using an alternative definition of long COVID (signs and symptoms at ≥ 4 weeks)

| | Overall cohort | | Immunocompetent cohort | | Immunocompromised cohort[1] | |
|---|---|---|---|---|---|---|
| | **Unvaccinated** (n=18,526) | **Primary series** (n=8,064) | **Unvaccinated** (n=16,864) | **Primary series** (n=7,972) | **Unvaccinated** (n=1,662) | **Primary series** (n=92) |
| Total number of primary care consultations | 56,428 | 19,036 | 50,593 | 18,716 | 5,835 | 320 |
| Mean follow-up, months (SD) | 3.1 (2.0) | 2.0 (1.3) | 3.1 (2.0) | 2.0 (1.3) | 3.1 (2.0) | 1.7 (0.9) |
| Crude person months | 57,343 | 16,467 | 52,121 | 16,306 | 5,221 | 161 |
| Unweighted primary care consultation rate per person months | 0.98 | 1.16 | 0.97 | 1.15 | 1.12 | 1.99 |
| Weighted primary care consultation rate per person months | 1.06 | 1.05 | 1.05 | 1.04 | - | - |
| Unweighted and unadjusted IRR (95% CI) | - | 1.17 (1.16-1.19)** | - | 1.18 (1.16-1.20)** | - | - |
| Weighted and adjusted IRR (95% CI) | - | 0.99 (0.96-1.03) | - | 0.99 (0.96-1.03) | - | - |

**p<0.01

Abbreviations: SD: standard deviation; IRR: incidence rate ratio; CI: confidence interval.

[1] Descriptive analyses are reported in the immunocompromised cohort due to the small sample size in the primary series group

**Supplementary table 7:** Primary care consultation rates and incidence rate ratio based on visits with a long COVID clinical diagnosis code, in long COVID patients, by immune system status

| | Overall cohort | | Immunocompetent cohort | | Immunocompromised cohort[1] | |
|---|---|---|---|---|---|---|
| | **Unvaccinated** (n=12,197) | **Primary series** (n=3,286) | **Unvaccinated** (n=11,079) | **Primary series** (n=3,250) | **Unvaccinated** (n=1,118) | **Primary series** (n=36) |
| Total number of primary care consultations | 17,629 | 4,114 | 15,885 | 4,067 | 1,744 | 47 |
| Mean follow-up, months (SD) | 2.3 (1.5) | 1.5 (1.1) | 2.3 (1.5) | 1.5 (1.1) | 2.4 (1.6) | 0.9 (0.6) |
| Crude person months | 28,642 | 4,889 | 25,989 | 4,856 | 2,653 | 33 |
| Unweighted primary care consultation rate per person months | 0.62 | 0.84 | 0.61 | 0.84 | 0.66 | 1.40 |
| Weighted primary care consultation rate per person months | 0.69 | 0.80 | 0.69 | 0.79 | - | - |
| Unweighted and unadjusted IRR (95% CI) | - | 1.37 (1.32-1.41)** | - | 1.37 (1.32-1.42)** | - | - |
| Weighted and adjusted IRR (95% CI) | - | 1.15 (1.11-1.21)** | - | 1.16 (1.11-1.21)** | - | - |

**p<0.01

Abbreviations: SD: standard deviation; IRR: incidence rate ratio; CI: confidence interval.

[1] Descriptive analyses are reported in the immunocompromised cohort due to the small sample size in the primary series group

### Supplementary table 8: Primary care consultation rates and incidence rate ratio in long COVID patients, by immune system status and high-risk status

**Patients at a higher risk of severe COVID-19**

| | **Overall high-risk cohort** | | **Patients at high-risk within the immunocompetent cohort** | |
|---|---|---|---|---|
| | Unvaccinated (n=2,036) | Primary series (n= 661) | Unvaccinated (n=1,735) | Primary series (n= 636) |
| Total number of primary care consultations | 6,764 | 1,493 | 5,729 | 1,418 |
| Mean follow-up, months (SD) | 2.4 (1.5) | 1.4 (1.1) | 2.4 (1.5) | 1.5 (1.1) |
| Crude person months | 4,846 | 958 | 4,094 | 933 |
| Unweighted primary care consultation rate per person months | 1.40 | 1.56 | 1.40 | 1.52 |
| Weighted primary care consultation rate per person months | 1.50 | 1.51 | 1.50 | 1.50 |
| Unweighted and unadjusted IRR (95% CI) | - | 1.12 (1.05-1.18)** | - | 1.09 (1.02-1.15)** |
| Weighted and adjusted IRR (95% CI) | - | 1.01 (0.91-1.11) | - | 1.00 (0.90-1.10) |

**Patients not at a higher risk of severe COVID-19**

| | **Patients not at high risk in the overall cohort** | | **Patients not at high-risk within the immunocompetent cohort** | |
|---|---|---|---|---|
| | Unvaccinated (n=10,161) | Primary series (n= 2,625) | Unvaccinated (n=9,344) | Primary series (n=2,614) |
| Total number of primary care consultations | 24,985 | 5,046 | 22,660 | 5,029 |
| Mean follow-up, months (SD) | 2.3 (1.5) | 1.5 (1.1) | 2.3 (1.5) | 1.5 (1.1) |
| Crude person months | 23,797 | 3,931 | 21,895 | 3,922 |
| Unweighted primary care consultation rate per 1,000 person months | 1.05 | 1.28 | 1.03 | 1.28 |
| Weighted primary care consultation rate per 1,000 person months | 1.15 | 1.18 | 1.14 | 1.18 |
| Unweighted and unadjusted IRR (95% CI) | - | 1.22 (1.19-1.26)** | - | 1.24 (1.20-1.28)** |
| Weighted and adjusted IRR (95% CI) | - | 1.02 (0.98-1.07) | - | 1.03 (0.98-1.08) |

**p<0.01

Abbreviations: SD: standard deviation; IRR: incidence rate ratio; CI: confidence interval.

**Supplementary table 9:** Primary care consultation costs in long COVID patients, based on visits with a long COVID clinical diagnosis code, by immune system

| | Overall cohort | | Immunocompetent cohort | | Immunocompromised cohort | |
|---|---|---|---|---|---|---|
| | **Unvaccinated** (n=12,197) | **Primary series** (n=3,286) | **Unvaccinated** (n=11,079) | **Primary series** (n=3,250) | **Unvaccinated** (n=1,118) | **Primary series** (n=36) |
| Mean primary care consultation costs, £ (SD) | 31.4 (33.0) | 27.4 (26.4) | 31.1 (32.7) | 27.4 (26.4) | 34.5 (35.8) | 28.6 (27.6) |
| Median primary care consultation costs, £ (Q1, Q3) | 15.5 (15.5, 39.2) | 15.5 (7.8, 39.2) | 15.5 (15.5, 39.2) | 15.5 (7.8, 39.2) | 23.4 (15.5, 47.0) | 15.5 (15.5, 42.9) |

Abbreviations: £: British pound sterling; SD: standard deviation; Q1: quartile 1; Q3: quartile 3.

Supplementary table 10: Primary care consultation costs in long COVID patients, by immune system, using an alternative definition of long COVID (signs and symptoms at ≥ 4 weeks)

| | **Overall cohort** | | **Immunocompetent cohort** | | **Immunocompromised cohort** | |
|---|---|---|---|---|---|---|
| | **Unvaccinated** n=18,526 | **Primary series** n=8,064 | **Unvaccinated** n=16,864 | **Primary series** n=7,972 | **Unvaccinated** n=1,662 | **Primary series** n=92 |
| Mean primary care consultation costs, £ (SD) | 41.5 (72.8) | 40.7 (56.8) | 40.6 (72.0) | 40.5 (56.3) | 49.8 (80.7) | 62.4 (87.2) |
| Median primary care consultation costs, £ (Q1, Q3) | 0 (0, 54.8) | 15.5 (0, 54.8) | 0 (0, 54.8) | 15.5 (0, 54.8) | 15.5 (0, 70.3) | 22.3 (0, 93.8) |

Abbreviations: £: British pound sterling; SD: standard deviation; Q1: quartile 1; Q3: quartile 3.

Supplementary file 1: Long COVID clinical code list. Long COVID symptom code lists are published by Subramanian, et al. 2022.

| Clinical Type | MedcodeID | SNOMED-CT code | Term |
|---|---|---|---|
| Diagnostic | 13486451000006115 | 1325161000000102 | Post-COVID-19 syndrome |
| Diagnostic | 13486491000006114 | 1325181000000106 | Ongoing symptomatic disease caused by severe acute respiratory syndrome coronavirus 2 |
| Diagnostic | | 1325181000000100 | Ongoing symptomatic COVID-19 |
| Diagnostic | 13486481000006111 | 1325181000000106 | Ongoing symptomatic COVID-19 |
| Diagnostic | 14168621000006112 | 1119303003 | Post-acute COVID-19 |
| Referral | 13486241000006118 | 1325021000000106 | Signposting to Your COVID Recovery |
| Referral | 13486251000006116 | 1325031000000108 | Referral to post-COVID assessment clinic |
| Referral | 13486261000006119 | 1325041000000104 | Referral to Your COVID Recovery rehabilitation platform |
| Referral | 13002611000006110 | 1321231000000101 | Signposting to CHMS (COVID-19 Home Management Service) |
| Referral | 14259201000006118 | 1325031000000108 | Referral to long term effects of COVID-19 assessment clinic |
| Assessment | 13486271000006114 | 1325051000000101 | Newcastle post-COVID syndrome Follow-up Screening Questionnaire |
| Assessment | 13486281000006112 | 1325061000000103 | Assessment using Newcastle post-COVID syndrome Follow-up Screening Questionnaire |
| Assessment | 13486301000006111 | 1325071000000105 | COVID-19 Yorkshire Rehabilitation Screening tool |
| Assessment | 13486311000006114 | 1325081000000107 | Assessment using COVID-19 Yorkshire Rehabilitation Screening tool |
| Assessment | 13486331000006115 | 1325091000000109 | Post-COVID-19 Functional Status Scale patient self-report |
| Assessment | 13486411000006116 | 1325101000000101 | Assessment using Post-COVID-19 Functional Status Scale patient self-report |
| Assessment | 13486371000006117 | 1325121000000105 | Post-COVID-19 Functional Status Scale patient self-report final scale grade |
| Assessment | 13486391000006116 | 1325131000000107 | Post-COVID-19 Functional Status Scale structured interview final scale grade |
| Assessment | 13486421000006112 | 1325141000000103 | Assessment using Post-COVID-19 Functional Status Scale structured interview |
| Assessment | 13486431000006110 | 1325151000000100 | Post-COVID-19 Functional Status Scale structured interview |

Supplementary file 2: Primary care unit costs obtained from the 2021 Personal Social Services Research Unit (PSSRU)

| | **Unit costs** |
|---|---|
| **Face-to-face consultations** | |
| General practitioner (GP) | £39.23 |
| Nurse | £6.76 |
| **Telephone consultations** | |
| GP | £15.32 |
| Nurse | £7.80 |
| **Online (e-)consultation** | £50.72 |
